# Respiratory microbiota signatures as a predictor of ventilator-associated pneumonia in hospitalised children with severe bronchiolitis

**DOI:** 10.64898/2026.01.13.26343576

**Authors:** Aleix Lluansí, Carmina Guitart, Miguel Blanco-Fuertes, Lluisa Hernández, Desirée Henares, Cèlia Martí-Castellote, Daniel Penela-Sanchez, Gerard González-Comino, Maria Cisneros, Mónica Balaguer, Carme Alejandre, Cristian Launes, Pedro Brotons, Carmen Muñoz-Almagro, Iolanda Jordan

## Abstract

**Objectives:** Ventilator-associated pneumonia (VAP) is a significant complication in pediatric patients with severe bronchiolitis undergoing mechanical ventilation (MV) in pediatric intensive care units (PICUs). The role of respiratory microbiota in VAP development remains underexplored in this vulnerable population. This study aimed to characterize respiratory microbiota in critically ill children with severe bronchiolitis receiving critical care and identify microbial patterns associated with VAP.

**Methods:** We conducted a cohort study in paediatric patients with severe bronchiolitis requiring MV at a tertiary PICU in Catalonia, Spain. Epidemiological, clinical and microbiological data were collected. Respiratory microbiota was assessed using 16S rRNA gene sequencing of nasopharyngeal (NP) aspirates and bronchoalveolar lavage (BAL) samples obtained before and during MV.

**Results:** Baseline NP microbiota differed significantly between VAP and non-VAP groups, with overrepresentation of *Moraxella, Enterobacter*, and *Amniculibacterium* genera and underrepresentation of *Prevotella* in patients who developed VAP. BAL microbiota showed fewer differences, although *Enterobacter* was more abundant in VAP cases. Random forest models demonstrated strong predictive performance, with the model integrating NP microbiota and clinical parameters achieving the highest accuracy (AUC 0.956).

**Conclusions:** Specific nasopharyngeal microbial signatures, combined with clinical factors, may serve as risk markers for VAP in mechanically ventilated children, potentially guiding targeted prevention strategies in PICUs.

## Introduction

Ventilator-associated pneumonia (VAP) is a significant complication in pediatric patients requiring mechanical ventilation (MV), particularly in those with severe bronchiolitis. Bronchiolitis is a common respiratory condition in infants and young children, primarily caused by viral infections, with respiratory syncytial virus (RSV) being the most prevalent pathogen [1]. About 10% of hospital admissions for RSV infection are severe enough to necessitate invasive ventilation in a Pediatric Intensive Care Unit (PICU) [2]. VAP is the second most common hospital-acquired infection in PICUs [3,4], with a prevalence of around 10-20% among pediatric patients who require MV [5–7]. VAP development has been associated with increased morbidity, mortality and healthcare costs. It often prolongs the duration of respiratory support and hospitalization, affecting patient outcomes in PICUs [8,9].

The pathogenesis of VAP in mechanically ventilated patients involves multiple factors, including micro-aspiration, biofilm formation within the endotracheal tube, the presence of bacteria in inhaled aerosols, translocation of bacteria from other niches, and in some cases, bloodstream infections [9,10]. Additionally, the role of the microbiome and host immune responses in VAP pathogenesis has been a growing area of research. The nasopharynx is a known reservoir for respiratory pathogens that can potentially migrate to the lower respiratory tract, especially in patients undergoing prolonged MV. However, the relationship between upper and lower respiratory microbiota and VAP has not been thoroughly explored.

The human respiratory tract harbors a diverse microbiota, which plays a crucial role in maintaining respiratory health and preventing infections. In particular, the nasopharyngeal (NP) microbiota serves as the first line of defense against invading pathogens [11–14]. However, in the context of MV and severe respiratory infections like bronchiolitis, this microbiota can be disrupted [15].

Longitudinal studies with daily cultures of endotracheal aspirates have suggested that the trachea is often colonized by causative pathogens at least 1-2 days before pneumonia develops, although not all colonizations progress to pneumonia [16]. Understanding the colonization of the upper airway and its potential progression to tracheobronchitis or VAP is critical. In addition, studies have suggested that certain microbial profiles in the upper respiratory tract could predispose patients to VAP [17], highlighting the need for further exploration of these microbial dynamics. Particularly, children with severe bronchiolitis who are admitted to PICUs represent a unique and vulnerable patient population, where the intersection of viral infection, immune response, the altered respiratory microbiota and the need for invasive respiratory support creates a high-risk environment for VAP development.

This study aimed to investigate the role of NP and bronchoalveolar lavage (BAL) microbiota in the development of VAP in hospitalized children with severe bronchiolitis admitted to the PICU, and to identify microbial baseline profiles associated with an increased risk of VAP that could help predicting its development.

## Materials and methods

### Study design

This cohort study included children under 24 months of age admitted to the PICU of a tertiary referral Children’s Hospital in Catalonia (Spain) for severe bronchiolitis, and who to required MV for more than 5 days. Participants were recruited between June 2020 and January 2023. The exclusion criteria were as follows: 1) clinical contraindication of BAL or broncoaspirate culture due to severe acute respiratory syndrome (PaO2/FiO2ratio<150); 2) pulmonary hemorrhage; 3) patients admitted to the PICU after MV for more than 24 hours in another hospital; 4) previous treatment with antibiotics (15 days before); 5) previous pneumonia (community or nosocomial); 6) and those that lacked parental consent.

### Definition of ventilator-associated pneumonia

VAP was defined by the following criteria: (1) PICU admission for more than 72 hours, and (2) MV via endotracheal intubation for a duration exceeding 48 hours. Diagnosis of VAP was established using the ENVIN-PED criteria, which are adapted from the Center for Disease Control (CDC) guidelines. These criteria encompass a combination of radiological, microbiological, and clinical-analytical indicators as outlined in the 2017 ENVIN-HELICS manual (available at: http://hws.vhebron.net/envin-helics/Help/Manual_2017.pdf).

### Sample and data collection

Epidemiological, clinical, and microbiological data were collected at intubation and at 3-5 days of MV: age, gender, weight, ethnicity, gestational age, delivery mode, feeding type, siblings, contact with pets, pathological antecedents, the pediatric risk of mortality (PRISM III) score [18], clinical pulmonary infection score (CPIS), leukocytes, C-reactive protein, procalcitonin, antibiotic during MV, bacterial cultures, and viral detection.

For microbiological analyses, nasopharyngeal aspirates (NPA) and BAL samples were collected at these two time points according to standard operational procedures at the study site. Samples were eluted in 2 ml of sterile PBS and frozen at –80°C in 1.5 mL DNa/RNase-free tubes.

### Bacterial cultures

Bacterial cultures were performed according to standard operational procedures and to the manufacturers’ instructions in the clinical laboratory of the setting. Requests for routine bacterial culture test were made at the discretion of clinicians based on patient presentation. Identification of bacterial species isolated by culture included Gram staining and matrix-assisted laser desorption/ionization time of flight mass spectrometry (MALDI-TOF).

### DNA extraction, viral RNA/DNA detection and bacterial sequencing

DNA was purified from NPA and BAL samples using the eMAG (bioMerieux; Marcy-l’Étoile, France) platform following the manufacturer’s instructions. A multiplex real-time PCR Anyplex II RV16 detection kit (Seegene) was used to test viral RNA/DNA from NPA samples. This system can detect and differentiate DNA/RNA from 16 of the most frequent human respiratory viruses: influenza A and B, respiratory syncytial virus A and B, adenovirus, enterovirus, parainfluenza virus 1, 2, 3 and 4, metapneumovirus, bocavirus, rhinovirus, coronavirus NL63, coronavirus 229E and coronavirus OC43. For bacterial sequencing, purified DNA samples were shipped on dry ice to Microomics Systems SL, Barcelona, Spain. The V3-V4 region of the bacterial 16S rRNA gene was amplified using previously described primers (V3–V4 16S rRNA gene region [19]). Additionally, negative controls (*i.e*., extraction and PCR) were included in the final library pool for controlling potential contamination (see details in Appendix 1).

### Bioinformatics analyses

All bioinformatics data analyses were conducted at the Microbial Genomics Unit of the Sant Joan de Deu Research Institute (IRSJD) using R software (v4.3.3, http://www.r-project.org/). The *DADA2* pipeline was followed to obtain exact amplicon sequence variants (ASVs) from raw reads [20]. Taxonomy at different levels was assigned to ASVs with 100% identity using the Ribosomal Database Project (RDP) version 11.5. The *Decontam* package was used for stricter contaminant elimination [21]. Samples were rarefied at 10,000 and 2,500 reads for NPA and BAL, respectively. A detailed description can be found in Appendix 1.

### Statistical analyses

Data description and differences on mean/medians or distribution was evaluated with standard statistical tests. Alpha diversity metrics (including Observed species, Chao1 richness, Shannon diversity and Inverse Simpson indexes) were calculated using the *Phyloseq* package [22]. For beta-diversity, a weighted UniFrac dissimilarity matrix was calculated on log-transformed relative abundance data. Differences in overall bacterial composition were tested with a nonparametric PERMANOVA test implemented in the Adonis2 function of the *vegan* package (v2.6-2), using 10,000 permutations. Differential abundances were tested at the genus taxonomic rank using Analysis of Compositions of Microbiomes with Bias Correction (ANCOMBC, v2.4.0). Three Random Forest (RF) classification models were built for predictive modelling using a 70/30 train-test split with 5-fold repeated cross-validation (5 repetitions) to ensure robust evaluation. The first model incorporated log-transformed abundance values from differential abundant genera identified by ANCOMBC. For the second model, the most predictive variable was selected from a broad set of epidemiological, clinical and microbiological data using Variable Selection Using Random Forests (*VSURF*) package [23]. The third model combined both the selected variable and differential abundant genera (see details in Appendix 1).

## Results

### Study population

A total of 35 children (females, n=16; 45.7%) were included in the study, of whom 8 developed VAP and 27 did not. The bacteria detected in BAL cultures for VAP confirmation in these patients are shown in Table S1. The median age was 1.4 months (IQR: 0.9-2.1 months), with no significant difference between the VAP (median: 1.2 months) and non-VAP (median: 1.4 months) groups (*p=*0.215). Median weight was 4.3 Kg (IQR: 3.3-5.9 Kg). No significant differences in demographic and epidemiological data were found between VAP and non-VAP groups (Table 1).

Pathologic health history was present in seven patients (20.0%), with similar proportions in the VAP (25.0%) and non-VAP (18.5%) groups. The PRISM III score was slightly higher (*p=*0.111) in the VAP group (median: 8.5, IQR: 1.8-10.3) than in the non-VAP group (median: 5.0, IQR: 0.0–6.0). Procalcitonin levels were significantly higher in VAP group compared to non-VAP (*p=*0.038). The use and duration of antibiotics were common (97.1%) and similar between both groups (median=7.0; *p*=0.681).

All patients were extracted NPA and BAL samples at both time points for bacterial culture. Notably, the detection of *Moraxella* in NPA cultures was significantly more frequent in VAP patients (*p*=0.010) but not in BAL samples (Table S2).

### Characterization of NP and BAL microbiota before and during MV

For NPA samples, all 35 patients’ samples were sequenced at time point 1. However, at time point 2, 29 NPA samples from these patients were sequenced, of which 24 met quality criteria for analyses (7 from the VAP group and 17 from the non-VAP group). For BAL samples, all were sequenced at both time points, but only 19 met the quality criteria at time point 1 (4 from the VAP group and 15 from the non-VAP group) and 20 at time point 2 (6 from the VAP group and 14 from the non-VAP group). Comparison of participants’ characteristics between VAP groups based on each time point and type of sample is depicted in Table S3.

A total of 2,034,401 denoised sequences from 59 NPA samples (median=15,726; IQR=5,962-33,331) were obtained, corresponding to 2,763 ASVs. For BAL samples, 791,812 denoised sequences from 39 samples (median=3,807; IQR=1,151-12,235) were obtained, corresponding to 7,069 ASVs.

Alpha diversity metrics showed no significant differences in NP bacterial richness (Observed and Chao1) or diversity (Shannon and Inverse Simpson) before and after MV (Figure S1A). However, the bacterial composition significantly differed before and at 3-5 days of MV (R^2^=8.2%, p*<*0.001, weighted UniFrac) (Figure S1B). In BAL samples, bacterial richness was significantly lower at 3-5 days of MV (Observed p<0.001; Chao1 p<0.001) (Figure S2A), although no significant differences in beta-diversity were observed (Adonis R2=6.5%, *p=*0.214) between the two time points (Figure S2B).

Taxonomic analyses identified 195 genera in NPA samples and 193 in BAL samples. However, only a subset of these genera (37 in NPA and 14 in BAL) had a prevalence higher than 10% across the samples. These prevalent genera accounted for approximately 94% of the total abundance detected in NPA (Table S4) and 87% in BAL samples (Table S5).The top-five most abundant genera in NPA samples at both time points included *Streptococcus* (22.3%), *Moraxella* (20.9%), *Haemophilus* (16.5%), *Prevotella* (6.6%), and *Staphylococcus* (4.1%) (Figure 1A, Table S4). In BAL samples, these same 5 genera (*Haemophilus* (32.5%), *Streptococcus* (19.0%), *Moraxella* (12.6%), *Staphylococcus* (9.0%), and *Prevotella* (2.2%) were predominantly observed, although additional genera such as *Enterobacter* and *Klebsiella* and *Serratia* were prominent in a few patients (Figure 1B, Table S5).

**Figure 1.**
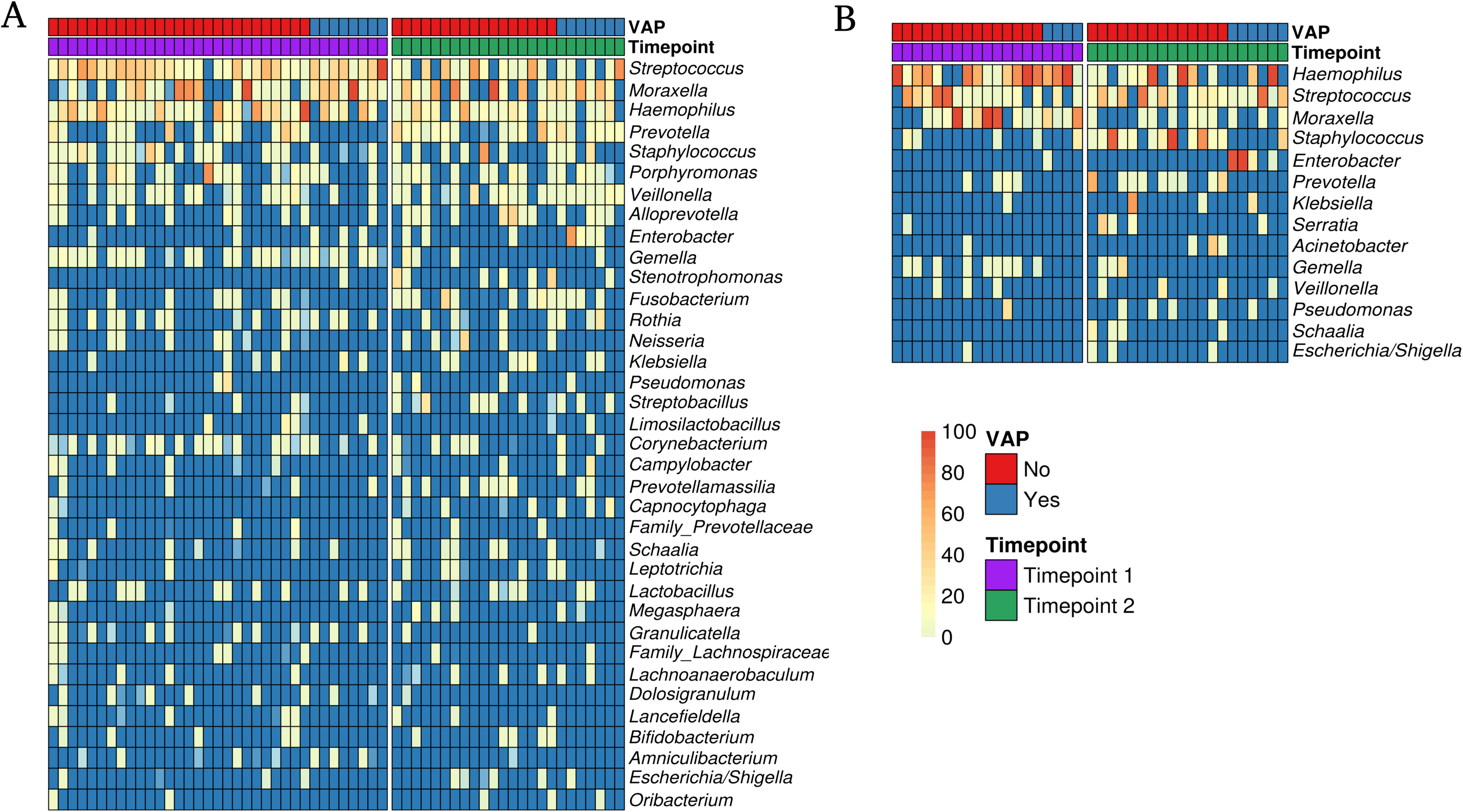
Characterization of nasopharyngeal (NP) and bronchoalveolar (BAL) microbiota in children with bronchiolitis prior and during mechanical ventilation (MV). Heatmap showing the relative abundance of main genera (prevalence >10%) in nasopharyngeal aspirate (NPA) (**A**) and bronchoalveolar lavage (BAL) (**B**) samples grouped by study time point (time point 1, pre-MV (n=35); time point 2, 3-5 days of MV (n=24)) and VAP development status. Genera are ordered based on their median relative abundance.

### Comparison of NP and BAL microbiota in children with bronchiolitis: VAP versus non-VAP

Comparative analyses of NP microbiota between the VAP and non-VAP groups at the two time points revealed similar alpha diversity metrics, with no significant differences in either bacterial richness or overall diversity (Figure 2A). However, significant differences in bacterial composition were observed prior to MV (Adonis R²=7.2%, *p=*0.032) (Figure 2B). Specifically, the VAP group exhibited an overrepresentation of *Moraxella*, *Enterobacter*, and *Amniculibacterium*, along with an underrepresentation of *Prevotella* at this initial time point. During the MV, although no significant changes in overall composition were found, a higher abundance of *Enterobacter* and *Veillonella* was detected in the VAP group (Figure 2C).

**Figure 2.**
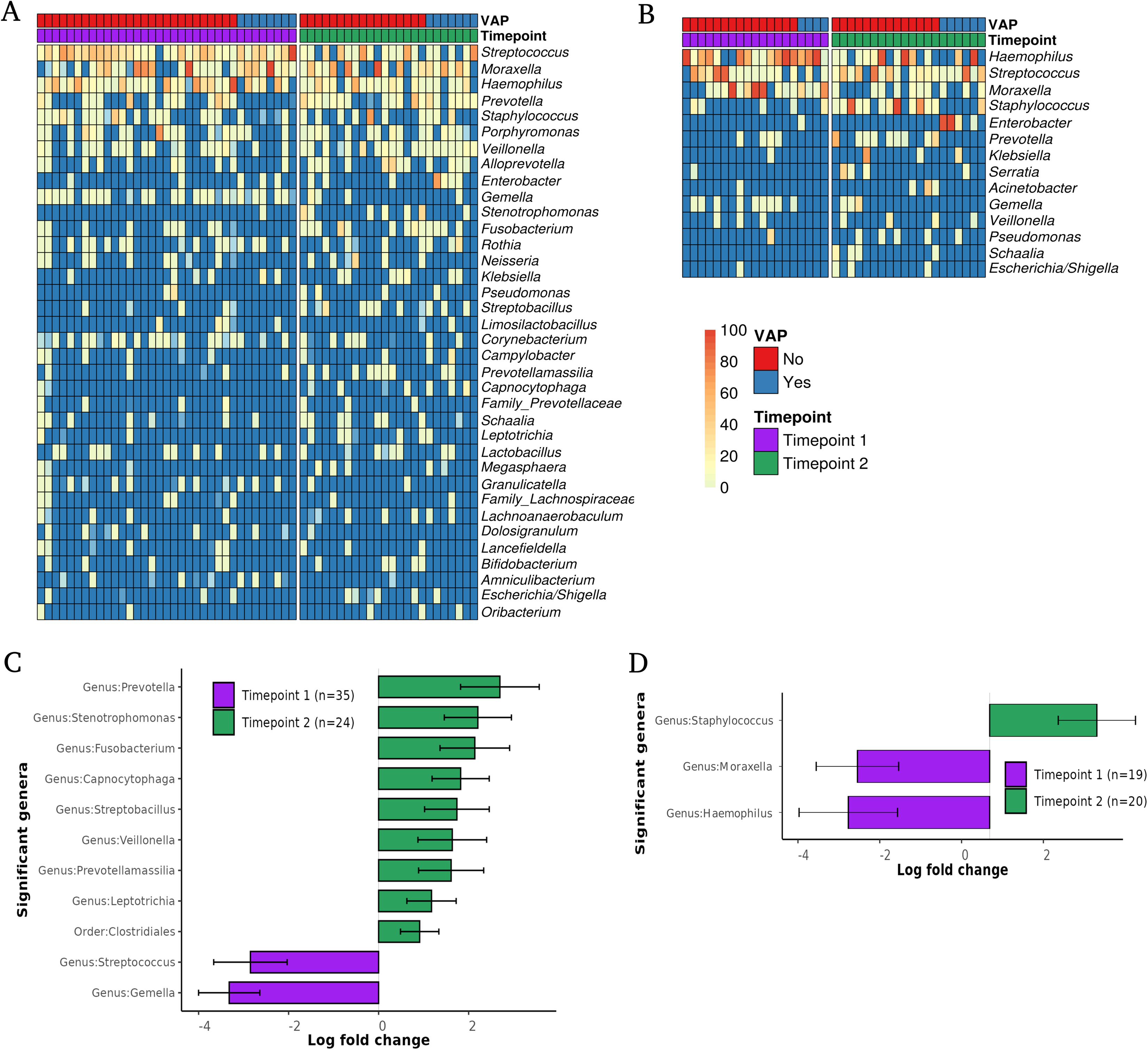
Comparison of nasopharyngeal (NP) microbiota between children with bronchiolitis who develop ventilator-associated pneumonia (VAP). (**A**) Boxplots with median and IQR comparing Observed species, Chao1, Shannon and Inverse Simpson indices of the NP microbiota at each study time point between VAP (pre-MV, n=8; 3-5 days of MV, n=7) and non-VAP (pre-MV, n=27; 3-5 days of MV, n=17) groups. P-values derived from standard statistical tests. (**B**) Principal Coordinate Analyses (PCoA) using weighted UniFrac dissimilarity matrix showing sample distribution at each study time point between VAP and non-VAP groups. Samples are connected to their corresponding centroids. P-value corresponds to Adonis PERMANOVA test. (**C**) Bar plots illustrating genera with significant differential abundance at each study time point between VAP and non-VAP groups, based on ANCOMBC analysis (p-value <0.05 and effect size > 2). Log fold changes are represented along the X-axis with standard error bars.

In the BAL microbiota, no significant differences in alpha diversity were detected between VAP groups prior to MV. However, a lower bacterial richness was detected at 3-5 days of MV (Chao1 *p=*0.015; Observed *p=*0.015) (Figure 3A), as well as suggestive differences in beta-diversity (*p=*0.057) (Figure 3B). In addition, a few differential abundant genera were identified (Figure 3C). Before MV, *Gemella* was more abundant in non-VAP group. At 3-5 days of MV, *Enterobacter* was overrepresented in VAP group, whereas *Staphylococcus* and *Prevotella* were more abundant in non-VAP group (Figure 3C).

**Figure 3.**
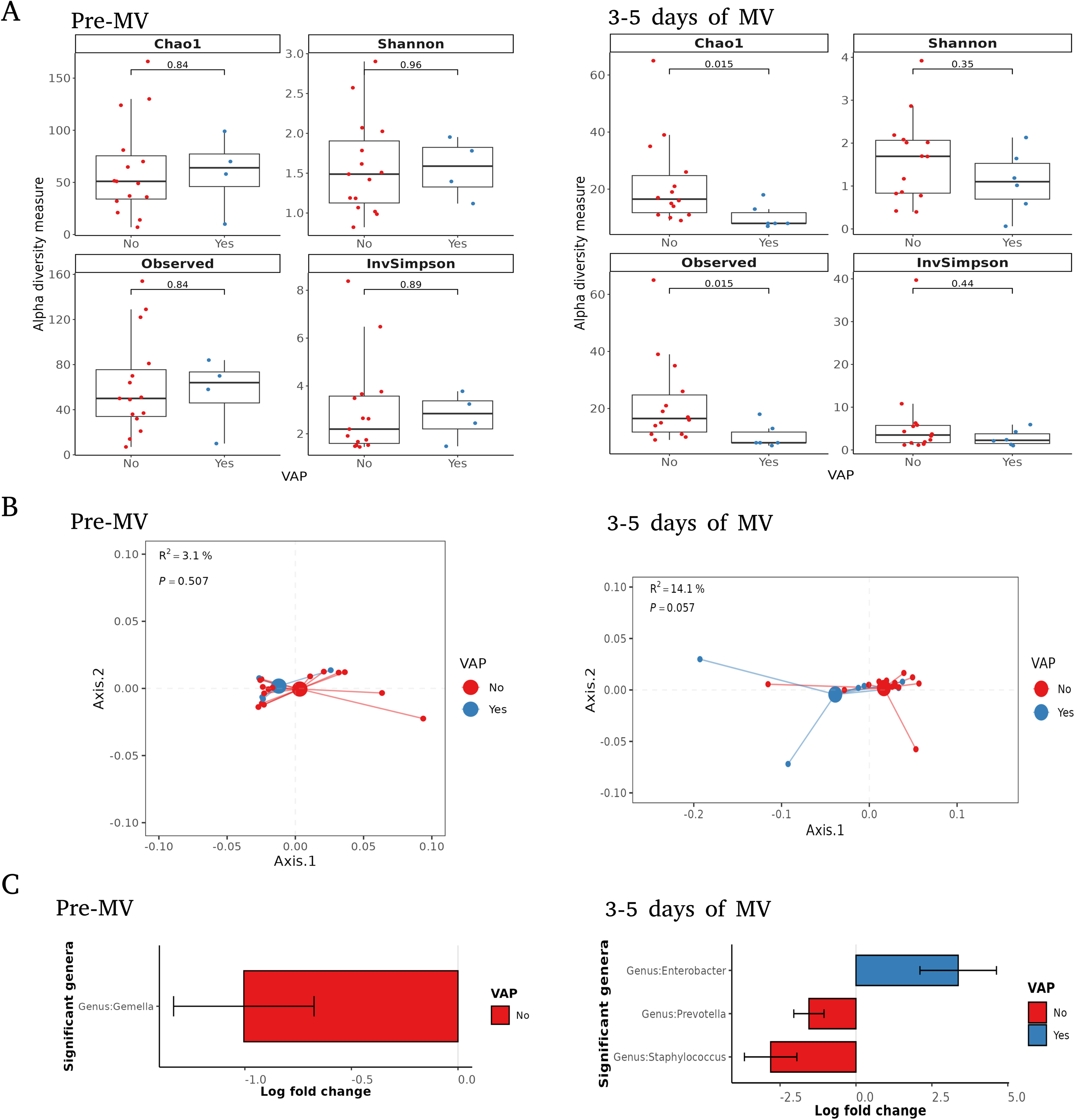
Comparison of bronchoalveolar lavage (BAL) microbiota between children with bronchiolitis who develop ventilator-associated pneumonia (VAP). (**A**) Boxplots with median and IQR comparing Observed species, Chao1, Shannon and Inverse Simpson indices of BAL microbiota at each study time point between VAP (pre-MV, n=4; 3-5 days of MV, n=6) and non-VAP (pre-MV, n=15; 3-5 days of MV, n=14) groups. P-values derived from standard statistical tests. (**B**) Principal Coordinate Analyses (PCoA) using weighted UniFrac dissimilarity matrix showing sample distribution at each study time point between VAP and non-VAP groups. Samples are connected to their corresponding centroids. P-value corresponds to Adonis PERMANOVA test. (**C**) Bar plots illustrating genera with significant differential abundance at each study time point between VAP and non-VAP groups, based on ANCOMBC analysis (p-value <0.05 and effect size > 2). Log fold changes are represented along the X-axis with standard error bars.

### Basal NP microbiota for VAP prediction using Random Forest

A RF model was developed to predict VAP in children with bronchiolitis, using the abundances of the four differentially abundant NP genera prior to MV (*i.e*., *Moraxella*, *Enterobacter*, *Amniculibacterium* and *Prevotella*). This model (Model 1) showed good predictive performance, obtaining a training AUC=0.781, a training accuracy=80%, and a testing accuracy=70%. The feature with the highest importance score in this model was the abundance of *Moraxella*. A second RF model (Model 2) was built incorporating the PRISM III score as it was identified as the most predictive variable by VSURF method. Model 2 achieved a training AUC=0.829, a training accuracy=80%, and a testing accuracy=80%. The third model (Model 3) combined the variables from Models 1 and 2, integrating NP genera abundances with the clinical predictor. This combined approach resulted in a training AUC of 0.956, a training accuracy of 92%, and a testing accuracy of 80%. In Model 3, PRISM III score was identified as the top feature with the highest importance score followed by *Enterobacter* and *Moraxella* abundance (Figure 4). Across all models, the features generally indicated an association with the VAP group. Of note, the abundance of *Prevotella* showed an inverse association with VAP in both Model 1 and Model 3 (Figure S3).

**Figure 4.**
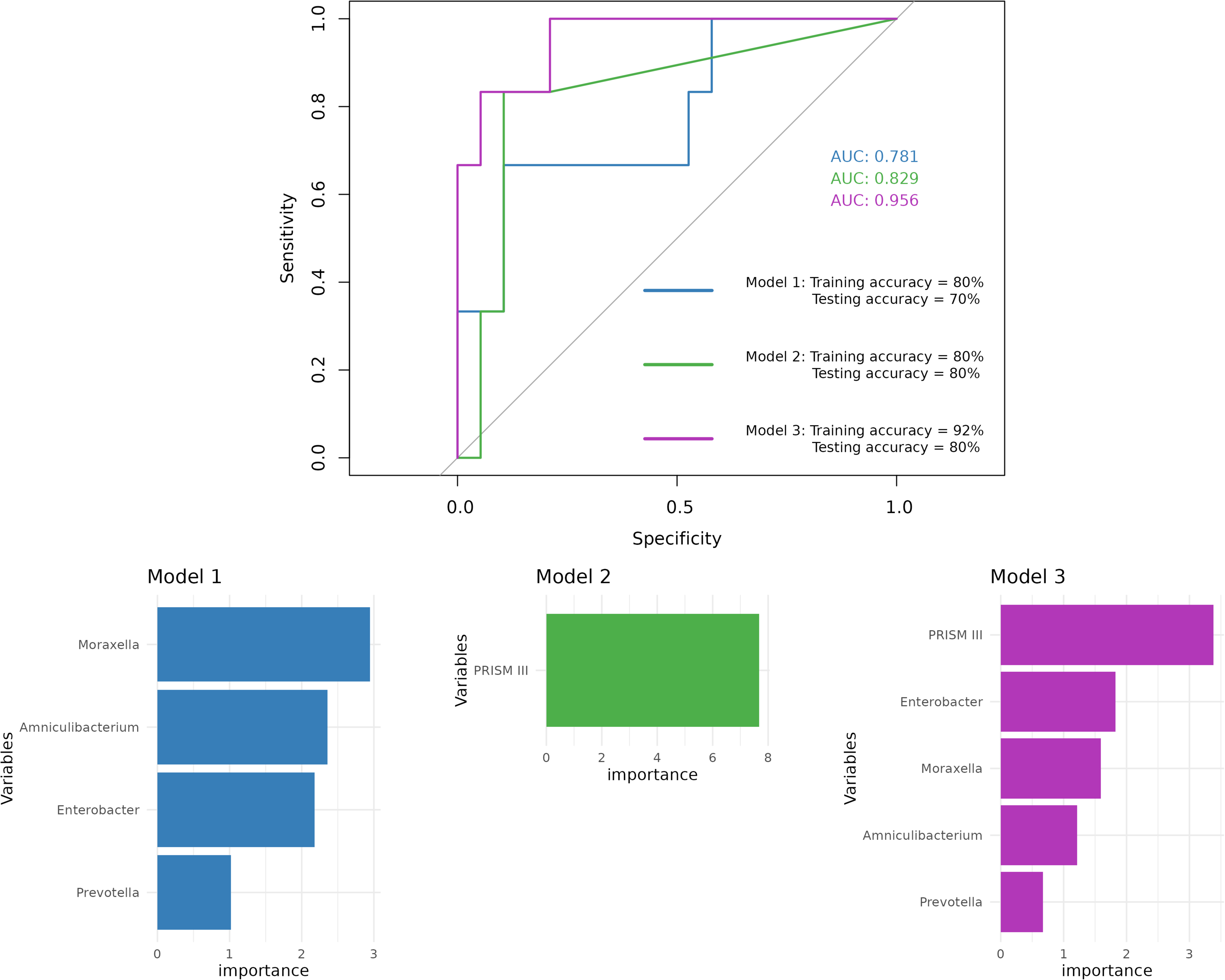
Random Forest models for predicting VAP development in children with severe bronchiolitis. (**A**) ROC curves of three RF models using 5-fold cross-validation with 5 repetitions for VAP prediction (n=35; 8 VAP, 27 non-VAP). Model 1 (blue) was built using the abundance values from identified significantly differential nasopharyngeal genera (*Moraxella*, *Enterobacter*, *Amniculibacterium* and *Prevotella*). Model 2 (green) included PRISM III score. Model 3 (purple) combined the variables from Model 1 and 2. (**B**) Bar plots displaying the Importance Score of each variable, representing their contribution to the performance of the three models.

## Discussion

The present study provides new insights into the role of NP and BAL microbiota in VAP development among pediatric patients with severe bronchiolitis admitted to a PICU. Our findings reveal distinct microbiota signatures that precede the development of VAP, notably the overrepresentation of genera such as *Moraxella* and *Enterobacter*, while highlighting the potential protective role of *Prevotella*. Through advanced predictive modelling, we demonstrate the feasibility of integrating microbial and clinical data to enhance early identification of VAP risk, offering promising avenues for targeted interventions in pediatric intensive care settings.

Our results showed no significant differences in overall alpha diversity metrics between VAP and non-VAP groups at both time points. However, the values observed, especially in NP samples, were lower than expected compared to studies of healthy children [14,24,25]. Previous studies have shown that a higher alpha diversity is typically associated with a healthy respiratory microbiota, which may provide resilience against colonization by potential pathogens [26–28]. Severe bronchiolitis, characterized by significant inflammation and airway obstruction, may disrupt the normal respiratory microbiota, leading to a state of low microbial richness and diversity and potentially contributing to a higher risk of subsequent infections such as VAP. In pediatric patients with bronchiolitis, there have been identified clusters of airway microbiota including a *Staphylococcus*-dominant profile and low proportions of *Moraxella, Corynebacterium* and *Dolosigranulum* [15].

In contrast to alpha diversity, significant differences in the bacterial composition of the NP microbiota were detected prior to MV, with specific genera being associated with a higher risk of developing VAP. Notably, children who developed VAP exhibited an overrepresentation of *Moraxella*, *Enterobacter*, and *Amniculibacterium* in the NP microbiota prior to MV, together with an underrepresentation of *Prevotella*. Some *Moraxella* species, such as *Moraxella catarrhalis*, are known respiratory pathogens capable of biofilm formation, which may enhance their persistence and promote VAP development [29]. *Enterobacter* species, which are frequently multidrug-resistant and associated with poor outcomes in PICUs, have been related to higher mortality in patients who develop VAP, predominantly in women [30]. Although *Amniculibacterium* is less commonly reported in VAP, it is phylogenetically very related to *Chryseobacterium* genus (16S rRNA gene sequence similarity of 91.8-96.0%) [31], which has been documented as a causative agent of VAP in immunocompromised patients, including infants and the elderly [32,33]. These findings align with previous studies suggesting that certain microbial profiles in the upper respiratory tract may increase susceptibility in VAP [34,35], highlighting the potential of specific microbial signatures as early markers of VAP risk. However, to our knowledge, this is the first study focused on children under 2 years of age suffering severe bronchiolitis.

The BAL microbiota revealed fewer differences in bacterial composition between the VAP and non-VAP groups at both time points, although significant decreases in bacterial richness were observed in the VAP group after 3–5 days of MV. This decrease in richness may reflect the selective pressure exerted by the hospital environment, antibiotics use, and the invasive nature of MV, which could promote the overrepresentation of nosocomial pathogens like *Enterobacter* in the lower respiratory tract. This genus was notably abundant in the VAP group’s BAL microbiota after 3–5 days on MV, whereas *Staphylococcus* and *Prevotella* were more abundant in the non-VAP group. The overrepresentation of *Enterobacter* in the VAP group is particularly concerning, as this organism is often multidrug-resistant and associated with poor outcomes in PICU patients [36–39].

Interestingly, in patients who developed VAP, *Enterobacter* was overrepresented both in the NP microbiota prior to MV and BAL during MV, but not in BAL prior to MV. This pattern suggests that *Enterobacter* may translocate from the upper to the lower respiratory tract during MV, potentially contributing to the development of VAP in these patients. The mechanisms facilitating this translocation could include micro-aspiration of oropharyngeal secretions, impaired mucociliary clearance due to endotracheal intubation, and the immunosuppressive effects associated with severe bronchiolitis and prolonged MV. These findings support the concept that the NP microbiota could act as a reservoir for pathogens that later colonize the lower airways, promoting VAP onset [16,40]. Thus, early detection of specific pathogens like *Enterobacter* in the upper airways might serve as a useful predictor for subsequent lower respiratory tract infections and a potential target for preventive interventions.

The inverse association of *Prevotella* with VAP risk observed in the basal NP microbiota, as indicated by RF models and differential abundance analysis, suggests that this genus may play a protective role, potentially by competing with or inhibiting the growth of pathogenic bacteria. In addition, results from BAL samples collected during the MV revealed an inverse association of this genus with VAP development. These findings are consistent with prior research suggesting that a higher abundance of certain commensal bacteria, such as some *Prevotella* species, may contribute to the maintenance of a balanced respiratory microbiome, thereby preventing colonization by more pathogenic bacteria [11,12,14]. Understanding the mechanisms underlying these protective effects could open new avenues for the prevention and management of VAP, such as the development of microbiota-modulating therapies.

The NP microbiota represents a less invasive method for sample collection than BAL, which could significantly enhance its utility in clinical practice. In this sense, our study also explored the potential of integrating basal NP microbial data and clinical parameters to develop predictive models for VAP in this population. The RF Model 3, which combined NP microbiota and clinical, provided the best performance with a training AUC of 0.956, a training accuracy of 92%, and a testing accuracy of 80%. Across all models, *Moraxella* abundance and PRISM III score were identified as the top features with the highest importance scores. The role of PRISM III role in quantifying illness severity in critically ill children with severe bronchiolitis might explain its presence as a top predictive variable in our VAP models. We observed a direct correlation of this score with VAP risk, although it did not reach statistical significance. Higher PRISM III scores indicate more severe physiological instability and often correspond with longer PICU stays and extended mechanical ventilation, both of which increase VAP susceptibility by compromising respiratory defenses. Additionally, PRISM III reflects immune function, with higher scores often indicating immune suppression that heightens vulnerability to pathogens like *Moraxella*, another key predictor in our models. Thus, PRISM III not only enhances predictive accuracy but also aligns with a multifaceted approach by providing insight into both physiological and immune factors that elevate VAP risk.

Despite its strengths, the study has some limitations. First, while the relatively small sample size, particularly after stratification by VAP status, may limit the generalizability of the findings, several measures were taken to ensure the robustness of the results. Cross-validation was employed, and the model was tested on 30% of the sample, providing support for the validity of the findings despite the sample size constraints. Nevertheless, the imbalance in group sizes could influence the performance of the RF models and warrants careful interpretation. Second, the observational nature of the study limits the ability to establish a definitive causal relationship between specific microbial profiles and VAP development. However, the longitudinal design strengthens the plausibility of such a relationship, which could be further confirmed with larger samples and interventional studies. Finally, while we identified microbial associations with VAP, the exact mechanisms by which these bacteria contribute to pathogenesis remain unclear. Further mechanistic studies are needed to elucidate these pathways.

In summary, our study provides evidence that specific nasopharyngeal microbial profiles, particularly the abundance of *Moraxella* and *Enterobacter* and the relative absence of *Prevotella*, are associated with an increased risk of VAP in children with severe bronchiolitis undergoing MV. The development of predictive models that integrate both microbial and clinical data holds promise for early identification of at-risk patients, potentially informing preventive strategies or guiding antibiotic use. These findings underscore the need for further research into the respiratory microbiome’s role in VAP pathogenesis and its potential as a target for preventative strategies in PICUs.

## Declarations

### Ethics approval and consent to participate

This research was approved by the Ethics Committee of Hospital Sant Joan de Déu (PIC-101-24) and was conducted in compliance with all relevant ethical regulations: the Helsinki Declaration of 1975 (revised in 2000) and the Spanish Organic Laws 15/1999 (December 13^th^) and 14/2007 (July 3^rd^) on data protection and biomedical research. All parents and/or legal guardians signed informed consent for the children to participate in the study.

### Consent for publication

All authors have consented to the publication of the results.

### Availability of data and materials

Sequence data that support the findings of this study have been deposited in the European Nucleotide Archive (ENA) at EMBL-EBI under primary accession number PRJEB83626. The associated clinical data that support the findings of this study are openly available at the CORA RDR repository at https://doi.org/10.34810/data1838

### Authors’ contribution

Conceptualization: C.M.-A., and I.J..; methodology: C.M.-A., I.J., and A.L..; recruitment and data collection: L.H., C.G, and C.M.-C.; resources: D.H., P.B., M.B.-F., D.P.-S., M.B., C.A., and C.L.; formal analyses: A.L.; data curation: A.L., L.H., C.G., and C.M.-C.; coordination: C.M.-A., and I.J.; writing of the manuscript: A.L.; revision of the manuscript: L.H.,C.G.,D.H., M.B.-F., P.B., D.P.-S.,G.G.-C.; C.M.-C., M.B., C.A., C.L., I.J., and C.M.-A. All authors read and approved the final manuscript.

## Acknowledgments

The authors acknowledge the support of all families who participated in this study. The authors would like to acknowledge Amaresh Pérez-Argüello for her contribution to microbiological studies. The results of this study were partially presented at the 10^th^ Congress of the European Academy of Paediatrics Society (EAPS) Vienna (Austria) 17-20 October 2024.

