## Supplementary material for "Respiratory microbiota signatures as a predictor of ventilator-associated pneumonia in hospitalised children with severe bronchiolitis"

**Supplementary tables**

**Table S1. Bacterial isolates identified in BAL cultures from VAP patients.**

| **VAP Patient** | **Bacterial Species Detected** |
| --- | --- |
| Patient 1 | Citrobacter koseri, Streptococcus pneumoniae |
| Patient 2 | Moraxella catarrhalis |
| Patient 3 | Enterobacter bugandensis, Haemophilus influenzae, Moraxella catarrhalis |
| Patient 4 | Moraxella catarrhalis |
| Patient 5 | Haemophilus influenzae, Staphylococcus aureus, Klebsiella varicola |
| Patient 6 | Serratia marcescens |
| Patient 7 | Haemophilus influenzae |
| Patient 8 | Moraxella catarrhalis, Staphylococcus aureus |

**Table S2. Comparison of microbiological variables between VAP and non-VAP groups at baseline.**

|  | **NPA samples** | | | **BAL samples** | | |
| --- | --- | --- | --- | --- | --- | --- |
| **Microbiological variables** | **VAP (n=8)** | **Non-VAP (n=27)** | **P-value** | **VAP (n=8)** | **Non-VAP (n=27)** | **P-value** |
| Bacterial detection in cultures, n (%) |  |  |  |  |  |  |
| Not done | 0 (0.0) | 2 (7.4) | 1.000 | 0 (0.0) | 1 (3.7) | 1.000 |
| *Haemophilus* detection | 1 (12.5) | 8 (29.6) | 0.608 | 0 (0.0) | 5 (18.5) | 0.460 |
| *Moraxella* detection | 6 (75.0) | 5 (18.5) | 0.010* | 3 (37.5) | 9 ( 33.3) | 1.000 |
| *Staphylococcus aureus* detection | 1 (12.5) | 4 (14.8) | 1.000 | 5 (62.5) | 9 ( 33.3) | 0.285 |
| *Streptococcus pneumoniae* detection | 1 (12.5) | 4 (14.8) | 1.000 | 1 (12.5) | 5 ( 18.5) | 1.000 |
| *Staphylococcus epidermidis* detection | 0 (0.0) | 0 (0.0) | 1.000 | 2 (25.0) | 5 (18.5) | 1.000 |
| *Streptococcus viridans* detection | 1 (12.5) | 0 (0.0) | 0.512 | 0 (0.0) | 0 (0.0) | 1.000 |
| *Escherichia coli* detection | 0 (0.0) | 1 (3.7) | 1.000 | 0 (0.0) | 0 (0.0) | 1.000 |
| *Klebsiella* detection | 1 (12.5) | 0 (0.0) | 0.512 | 0 (0.0) | 0 (0.0) | 1.000 |
| *Serratia* detection | 0 (0.0) | 0 (0.0) | 1.000 | 1 (12.5) | 1 (3.7) | 0.941 |
| *Pseudomonas* detection | 0 (0.0) | 1 (3.7) | 1.000 | 0 (0.0) | 1 (3.7) | 1.000 |
| *Enterobacter* detection | 1 (12.5) | 0 (0.0) | 0.512 | 0 (0.0) | 1 (3.7) | 1.000 |
| *Bordetella* detection | 0 (0.0) | 0 (0.0) | 1.000 | 1 (12.5) | 0 (0.0) | 0.512 |
| *Citrobacter* detection | 1 (12.5) | 0 (0.0) | 0.512 | 1 (12.5) | 0 (0.0) | 0.512 |
| *Streptococcus agalactiae* detection | 0 (0.0) | 1 (3.7) | 1.000 | 0 (0.0) | 1 (3.7) | 1.000 |
| Other bacteria detected | 0 (0.0) | 1 (3.7) | 1.000 | 0 (0.0) | 1 (3.7) | 1.000 |
| No bacteria detected | 0 (0.0) | 6 (22.2) | 0.352 | 0 (0.0) | 0 (0.0) | 1.000 |
| Viral detection, n (%) | 8/8 (100.0) | 18/20 (90.0) | 0.907 | ND | ND | ND |

Proportions between the groups were compared using Pearson Chi-square test. *p-value ≤ 0.05. ND: not determined.

**Table 2. Comparison of participants’ characteristics according to VAP development status for each type of sample and study timepoint.**

|  | **Timepoint 1 (pre-MV)** | | | | | | **Timepoint 2 (3-5 days of MV)** | | | | | |
| --- | --- | --- | --- | --- | --- | --- | --- | --- | --- | --- | --- | --- |
|  | **NPA samples** | | | **BAL samples** | | | **NPA samples** | | | **BAL samples** | | |
|  | **VAP**  **(n=8)** | **Non-VAP (n=27)** | **P-value** | **VAP**  **(n=4)** | **Non-VAP (n=15)** | **P-value** | **VAP**  **(n=7)** | **Non-VAP (n=17)** | **P-value** | **VAP**  **(n=6)** | **Non-VAP (n=14)** | **P-value** |
| **Demographic and epidemiological variables** |  |  |  |  |  |  |  |  |  |  |  |  |
| Age, months (median, IQR) | 1.2 (0.7-1.8) | 1.4 (1.0-2.6) | 0.215 | 1.1 (0.7-1.6) | 1.4 (1.0-2.1) | 0.394 | 0.9 (0.7-1.8) | 1.4 (1.0-1.8) | 0.293 | 1.1 (0.8-1.6) | 1.1 (0.9-2.0) | 0.535 |
| Sex, female (%) | 2 (25.0) | 14 (51.9) | 0.350 | 1 (25.0) | 6 (40.0) | 1 | 2 (28.6) | 9 (52.9) | 0.523 | 1 (16.7) | 4 (28.6) | 1 |
| Weight, Kg (median, IQR) | 3.35 (3.2-4.2) | 4.7 (3.9-6.0) | 0.070 | 3.3 (3.2-4.0) | 4.9 (3.8-6.0) | 0.246 | 3.3 (3.0-3.5) | 4.6 (3.7-6.0) | 0.075 | 3.3 (3.2-3.5) | 4.6 (4.0-5.9) | 0.107 |
| Ethnicity, aucasian, n (%) | 7 (87.5) | 23 (85.2) | 0.856 | 4 (100.0) | 13 (86.7) | 0.742 | 6 (85.7) | 14 (82.4) | 0.801 | 5 (83.3) | 13 (92.9) | 0.246 |
| Median gestational age, weeks (median, IQR) | 37.5 (36.5-39.0) | 39.0 (36.0-39.7) | 0.446 | 37.0 (35.7-37.5) | 38.0 (36.5-40.0) | 0.266 | 37 (36-39) | 39 (35-40) | 0.585 | 37.0 (35.5-37.7) | 38.5 (36.2-39.7) | 0.197 |
| Delivery mode, C-section (%) | 3 (37.5) | 4 (14.8) | 0.365 | 2 (50.0) | 1 (6.7) | 0.180 | 3 (42.8) | 3 (17.6) | 0.437 | 3 (50.0) | 4 (28.6) | 0.682 |
| Feeding type, n (%) |  |  |  |  |  |  |  |  |  |  |  |  |
| Breastfeeding | 1 (12.5) | 8/26 (30.7) | 0.571 | 0 (0.0) | 4 (26.7) | 0.391 | 0 (0.0) | 4/16 (25.0) | 0.637 | 1 (16.7) | 3 (21.4) | 1 |
| Formula-feeding | 5 (62.5) | 12/26 (46.1) | 0.686 | 3 (75.0) | 8 (53.3) | 0.619 | 5 (71.4) | 8/16 (50.0) | 0.833 | 4 (66.6) | 6 (42.9) | 0.735 |
| Mixed formula and breast milk | 2 (25.0) | 6/26 (23.1) | 1 | 1 (25.0) | 3 (20.0) | 1 | 2 (28.6) | 2/16 (12.5) | 1 | 1 (16.7) | 4 (28.6) | 0.929 |
| Siblings, n (%) | 8 (100.0) | 22 (81.5) | 0.580 | 4 (100.0) | 15 (100.0) | 1 | 7 (100.0) | 13/16 (81.3) | 0.578 | 6 (100.0) | 11/13 (84.6) | 0.832 |
| Contact with pets, n (%) | 1/4 (25.0) | 6/18 (33.3) | 1 | 1 (25.0) | 4 (26.7) | 1 | 1/4 (25.0) | 4/12 (33.3) | 1 | 1/2 (50.0) | 5/6 (83.3) | 1 |
| **Clinical variables** |  |  |  |  |  |  |  |  |  |  |  |  |
| Pathologic antecedents, n (%) | 2 (25.0) | 5 (18.5) | 1 | 1 (25.0) | 3 (20.0) | 1 | 2 (28.6) | 3 (17.6) | 0.963 | 2 (33.3) | 4 (28.6) | 1 |
| PRISM III (median, IQR) | 8.5 (1.8-10.3) | 5.0 (0.0-6.0) | 0.111 | 9.0 (6.5-10.7) | 5.0 (1.5-6.0) | 0.068 | 9.0 (5.0-10.5) | 5.0 (3.0-6.0) | 0.084 | 5.0 (1.0-10.2) | 5.0 (0.7-6.0) | 0.450 |
| CPIS (median, IQR) | 6.5 (5.3-8.5) | NA | NA | 7.0 (6.0-8.0) | NA | NA | 6.5 (5.3-8.5) | NA | NA | 6.0 (5.2-8.5) | NA | NA |
| Leukocytes, thousandmm^−3^ (mean/median, SD/IQR) | 9062.5 (6045.3) | 9330.1 (5028.6) | 0.911 | 13075.0 (6374.1) | 8573.3 (4162.7) | 0.293 | 9600.0 (6319.0) | 9236.1 (5785.6) | 0.901 | 8683.3 (6771.2) | 7672.4 (5395.4) | 0.754 |
| C-reactive protein, mg/dL (mean, SD) | 120.9 (59.9) | 83.1 (58.0) | 0.142 | 124.6 (45.4) | 92.7 (53.5) | 0.279 | 109.7 (54.8) | 96.5 (63.3) | 0.664 | 130.9 (63.5) | 74.4 (60.7) | 0.098 |
| Procalcitonin, ng/mL (median, IQR) | 7.1 (0.8-18.9) | 0.9 (0.3-2.6) | 0.038* | 19.8 (13.7-30.1) | 0.9 (0.3-2.5) | 0.061 | 7.5 (0.7-19.8) | 1.3 (0.4-2.7) | 0.193 | 12.7 (6.8-20.7) | 0.4 (0.2-2.7) | 0.002* |
| Antibiotic during MV, n (%) |  |  |  |  |  |  |  |  |  |  |  |  |
| Betalactam | NA | NA | NA | NA | NA | NA | 6 (85.7) | 17 (100.0) | 1 | 6 (100.0) | 13 (92.9) | 0.513 |
| Non-betalactam | NA | NA | NA | NA | NA | NA | 2 (28.6) | 0 (0.0) | 0.188 | 4 (66.6) | 3 (21.4) | 0.266 |

^α^ Data from breastfed and mixed milk fed infants.

Proportions between the groups were compared using Pearson’s Chi-square test. For continuous variables, the t-test for normally distributed data and the Wilcoxon Rank Sum test for non-normally distributed data were performed. *p-value ≤ 0.05.

IQR: interquartile range; VAP: ventilator-associated pneumonia; SD: standard deviation; MV: mechanical ventilation; PRISM III: paediatric risk of mortality score; NPA: nasopharyngeal aspirate; BAL: bronchoalveolar lavage; CPIS: clinical pulmonary infection score; NA: not applicable

**Table S4. Relative abundance of genera with a prevalence greater than 10% across in NPA samples**

| **Kingdom** | **Phylum** | **Class** | **Order** | **Family** | **Genus** | **Mean ± SD relative abundance (%)** |
| --- | --- | --- | --- | --- | --- | --- |
| Bacteria | Firmicutes | Bacilli | Lactobacillales | Streptococcaceae | Streptococcus | 22.27 ± 22.62 |
| Bacteria | Proteobacteria | Gammaproteobacteria | Pseudomonadales | Moraxellaceae | Moraxella | 20.85 ± 28.77 |
| Bacteria | Proteobacteria | Gammaproteobacteria | Pasteurellales | Pasteurellaceae | Haemophilus | 16.49 ± 21.13 |
| Bacteria | Bacteroidetes | Bacteroidia | Bacteroidales | Prevotellaceae | Prevotella | 6.64 ± 11.88 |
| Bacteria | Firmicutes | Bacilli | Bacillales | Staphylococcaceae | Staphylococcus | 4.14 ± 11.26 |
| Bacteria | Bacteroidetes | Bacteroidia | Bacteroidales | Porphyromonadaceae | Porphyromonas | 2.89 ± 5.77 |
| Bacteria | Bacteroidetes | Bacteroidia | Bacteroidales | Prevotellaceae | Alloprevotella | 2.49 ± 6.66 |
| Bacteria | Firmicutes | Negativicutes | Veillonellales | Veillonellaceae | Veillonella | 2.42 ± 5.11 |
| Bacteria | Proteobacteria | Gammaproteobacteria | Enterobacterales | Enterobacteriaceae | Enterobacter | 1.74 ± 9.33 |
| Bacteria | Firmicutes | Bacilli | Bacillales | Bacillales_Incertae_Sedis_XI | Gemella | 1.55 ± 3.01 |
| Bacteria | Proteobacteria | Gammaproteobacteria | Xanthomonadales | Xanthomonadaceae | Stenotrophomonas | 1.45 ± 5.93 |
| Bacteria | Fusobacteria | Fusobacteriia | Fusobacteriales | Fusobacteriaceae | Fusobacterium | 1.39 ± 5.11 |
| Bacteria | Actinobacteria | Actinobacteria | Micrococcales | Micrococcaceae | Rothia | 1.26 ± 3.64 |
| Bacteria | Proteobacteria | Betaproteobacteria | Neisseriales | Neisseriaceae | Neisseria | 1.01 ± 4.83 |
| Bacteria | Proteobacteria | Gammaproteobacteria | Enterobacterales | Enterobacteriaceae | Klebsiella | 0.81 ± 2.94 |
| Bacteria | Proteobacteria | Gammaproteobacteria | Pseudomonadales | Pseudomonadaceae | Pseudomonas | 0.76 ± 3.98 |
| Bacteria | Fusobacteria | Fusobacteriia | Fusobacteriales | Leptotrichiaceae | Streptobacillus | 0.63 ± 2.95 |
| Bacteria | Actinobacteria | Actinobacteria | Mycobacteriales | Corynebacteriaceae | Corynebacterium | 0.56 ± 1.66 |
| Bacteria | Campilobacterota | Campylobacteria | Campylobacterales | Campylobacteraceae | Campylobacter | 0.53 ± 3.22 |
| Bacteria | Bacteroidetes | Flavobacteriia | Flavobacteriales | Flavobacteriaceae | Capnocytophaga | 0.49 ± 2.55 |
| Bacteria | Bacteroidetes | Bacteroidia | Bacteroidales | Prevotellaceae | Prevotellamassilia | 0.48 ± 1.48 |
| Bacteria | Firmicutes | Bacilli | Lactobacillales | Lactobacillaceae | Limosilactobacillus | 0.37 ± 2.21 |
| Bacteria | Bacteroidetes | Bacteroidia | Bacteroidales | Prevotellaceae | NA | 0.32 ± 1.28 |
| Bacteria | Actinobacteria | Actinobacteria | Actinomycetales | Actinomycetaceae | Schaalia | 0.28 ± 0.88 |
| Bacteria | Fusobacteria | Fusobacteriia | Fusobacteriales | Leptotrichiaceae | Leptotrichia | 0.28 ± 1.25 |
| Bacteria | Firmicutes | Bacilli | Lactobacillales | Lactobacillaceae | Lactobacillus | 0.22 ± 0.64 |
| Bacteria | Firmicutes | Bacilli | Lactobacillales | Carnobacteriaceae | Granulicatella | 0.20 ± 0.54 |
| Bacteria | Firmicutes | Negativicutes | Veillonellales | Veillonellaceae | Megasphaera | 0.20 ± 1.17 |
| Bacteria | Firmicutes | Clostridia | Clostridiales | Lachnospiraceae | NA | 0.17 ± 0.70 |
| Bacteria | Firmicutes | Clostridia | Clostridiales | Lachnospiraceae | Lachnoanaerobaculum | 0.14 ± 0.47 |
| Bacteria | Firmicutes | Bacilli | Lactobacillales | Carnobacteriaceae | Dolosigranulum | 0.13 ± 0.70 |
| Bacteria | Actinobacteria | Actinobacteria | Bifidobacteriales | Bifidobacteriaceae | Bifidobacterium | 0.11 ± 0.35 |
| Bacteria | Actinobacteria | Coriobacteriia | Coriobacteriales | Atopobiaceae | Lancefieldella | 0.11 ± 0.39 |
| Bacteria | Bacteroidetes | Flavobacteriia | Flavobacteriales | Flavobacteriaceae | Amniculibacterium | 0.10 ± 0.36 |
| Bacteria | Proteobacteria | Gammaproteobacteria | Enterobacterales | Enterobacteriaceae | Escherichia/Shigella | 0.02 ± 0.07 |

SD: Standard deviation

**Table S5. Relative abundance of genera with a prevalence greater than 10% across in BAL samples.**

| **Kingdom** | **Phylum** | **Class** | **Order** | **Family** | **Genus** | **Mean ± SD relative abundance (%)** |
| --- | --- | --- | --- | --- | --- | --- |
| Bacteria | Proteobacteria | Gammaproteobacteria | Pasteurellales | Pasteurellaceae | Haemophilus | 32.45 ± 39.72 |
| Bacteria | Firmicutes | Bacilli | Lactobacillales | Streptococcaceae | Streptococcus | 19.01 ± 28.57 |
| Bacteria | Proteobacteria | Gammaproteobacteria | Pseudomonadales | Moraxellaceae | Moraxella | 12.60 ± 28.30 |
| Bacteria | Firmicutes | Bacilli | Bacillales | Staphylococcaceae | Staphylococcus | 9.06 ± 24.00 |
| Bacteria | Proteobacteria | Gammaproteobacteria | Enterobacterales | Enterobacteriaceae | Enterobacter | 4.43 ± 21.73 |
| Bacteria | Bacteroidetes | Bacteroidia | Bacteroidales | Prevotellaceae | Prevotella | 2.15 ± 11.22 |
| Bacteria | Proteobacteria | Gammaproteobacteria | Enterobacterales | Enterobacteriaceae | Klebsiella | 2.06 ± 11.86 |
| Bacteria | Proteobacteria | Gammaproteobacteria | Enterobacterales | Yersiniaceae | Serratia | 1.45 ± 6.51 |
| Bacteria | Firmicutes | Bacilli | Bacillales | Bacillales_Incertae_Sedis_XI | Gemella | 1.06 ± 5.32 |
| Bacteria | Proteobacteria | Gammaproteobacteria | Pseudomonadales | Pseudomonadaceae | Pseudomonas | 0.82 ± 4.95 |
| Bacteria | Proteobacteria | Gammaproteobacteria | Pseudomonadales | Moraxellaceae | Acinetobacter | 0.80 ± 6.87 |
| Bacteria | Firmicutes | Negativicutes | Veillonellales | Veillonellaceae | Veillonella | 0.63 ± 4.63 |
| Bacteria | Actinobacteria | Actinobacteria | Actinomycetales | Actinomycetaceae | Schaalia | 0.45 ± 3.07 |
| Bacteria | Proteobacteria | Gammaproteobacteria | Enterobacterales | Enterobacteriaceae | Escherichia/Shigella | 0.12 ± 0.54 |

SD: Standard deviation

**Supplementary figures**

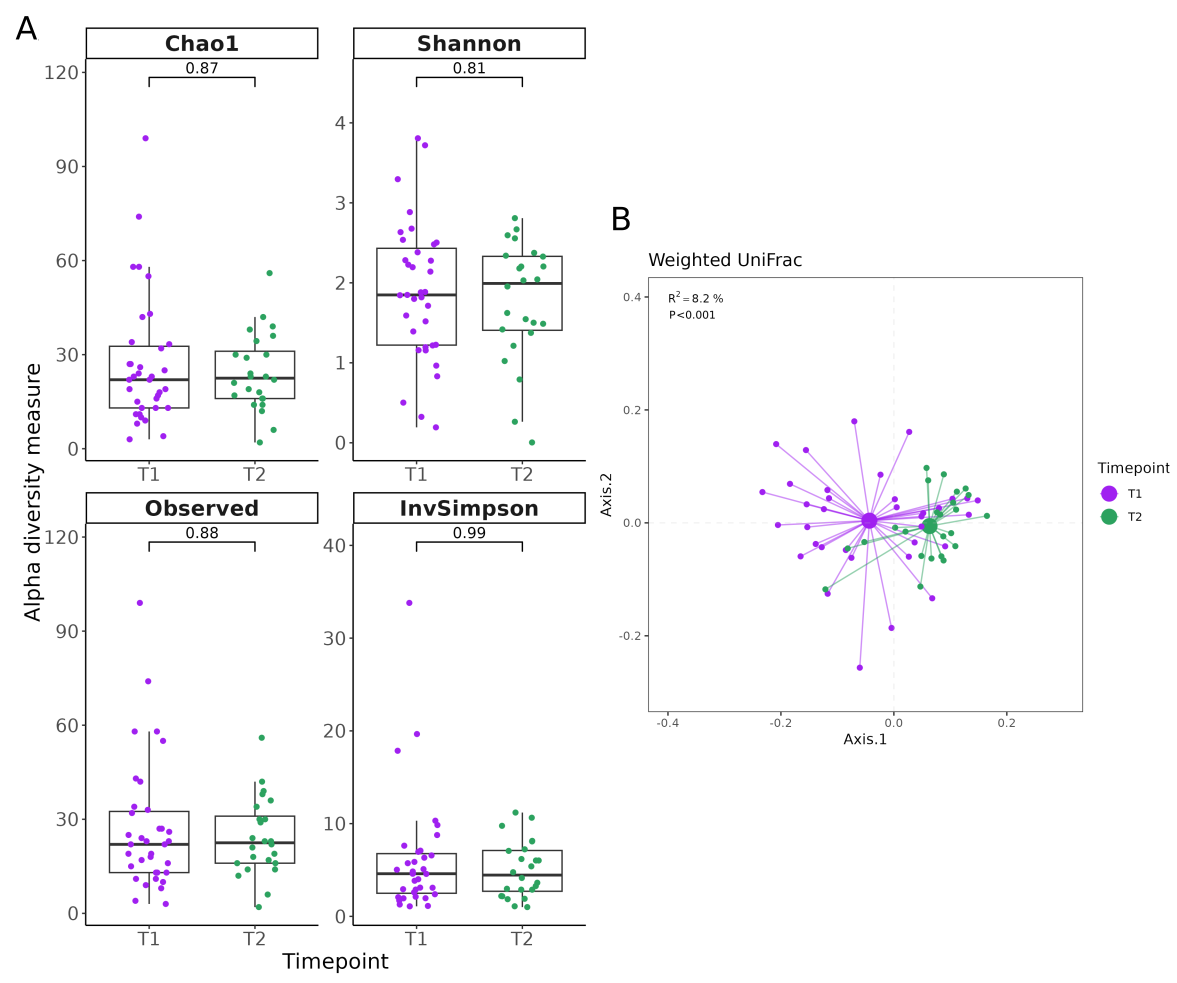

**Figure S1. Differences in alpha- and beta-diversity between study time points in nasopharyngeal aspirate (NPA) samples. (A**) Comparison of the Chao1, Shannon, Observed OTUs, and Inversed Simpson indices for bacterial richness and diversity. The boxes show the medians and interquartile ranges, and the whiskers indicate the 5th to 95th percentiles. (**B**) Principal coordinate analysis (PCoA) of the nasopharyngeal microbiota with samples clustered by time points and connected with their corresponding centroid. Represented distances are based on weighted UniFrac.

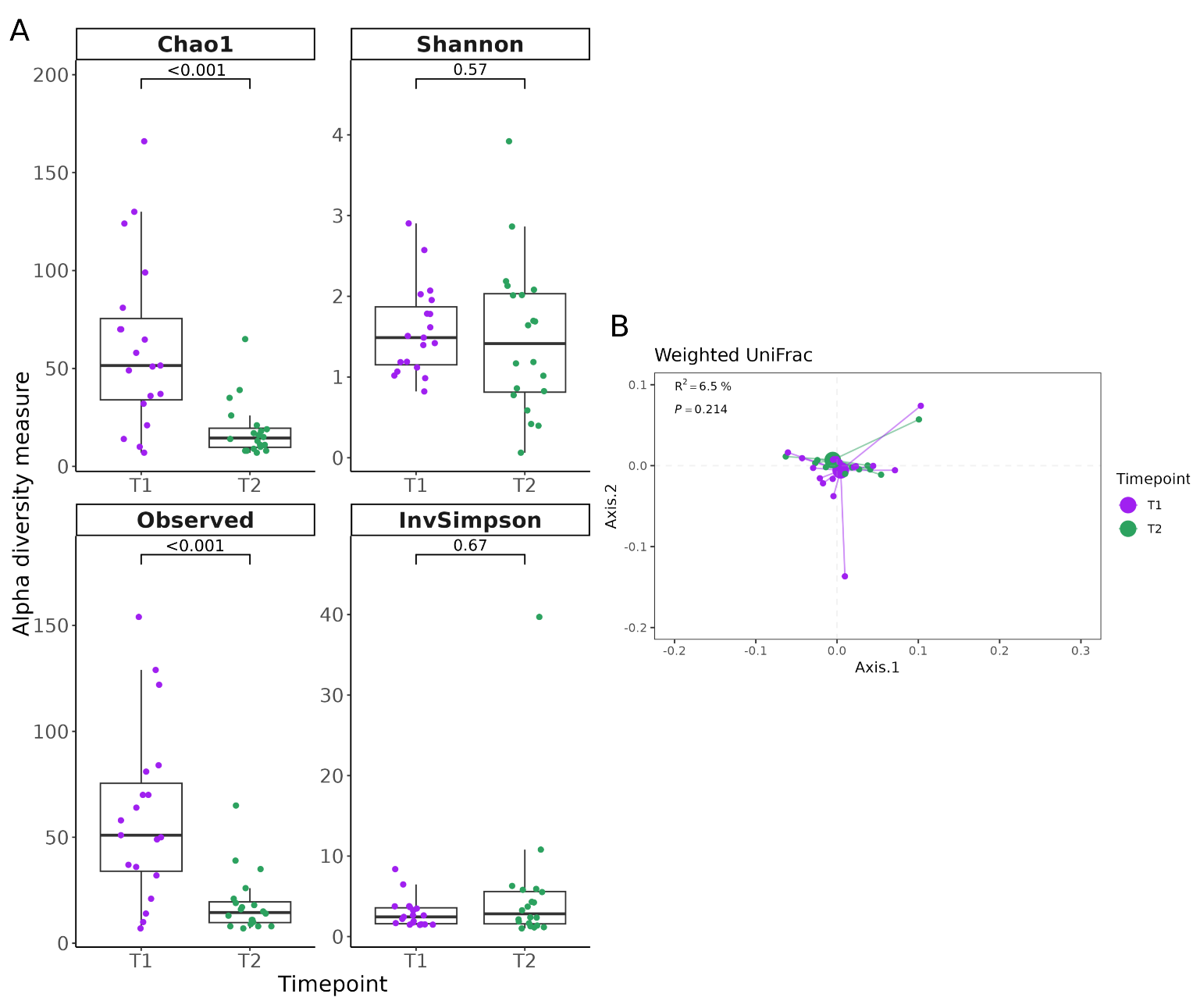

**Figure S2. Differences in alpha- and beta-diversity between study time points in bronchoalveolar lavage (BAL) samples. (A**) Comparison of the Chao1, Shannon, Observed OTUs, and Inversed Simpson indices for bacterial richness and diversity. The boxes show the medians and interquartile ranges, and the whiskers indicate the 5th to 95th percentiles. (**B**) Principal coordinate analysis (PCoA) of the BAL microbiota with samples clustered by time points and connected with their corresponding centroid. Represented distances are based on weighted UniFrac.

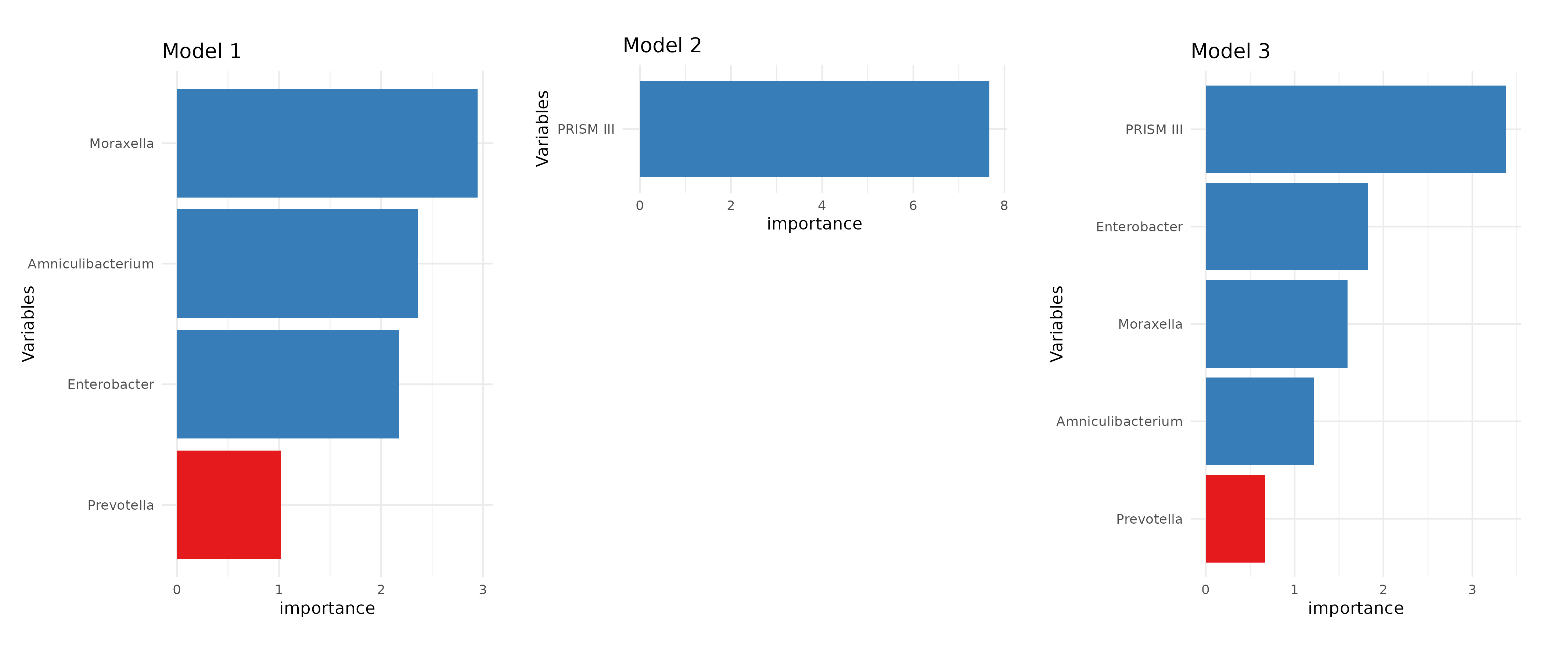

**Figure S3. Contribution of Variables to Model Performance.** Bar plots display the mean decrease in the Gini index for each variable, indicating its importance in the performance of the three models. The colour coding shows the direction of the association: blue represents variables associated with ventilator-associated pneumonia (VAP), while red represents variables associated with non-VAP.
